# Navigating life with HIV as an older adult on the Kenyan coast: perceived health challenges seen through the biopsychosocial model

**DOI:** 10.1101/2022.02.27.22271072

**Authors:** Patrick N Mwangala, Ryan G Wagner, Charles R Newton, Amina Abubakar

**Affiliations:** Centre for Geographic Medicine Research Coast, Kenya Medical Research Institute (KEMRI), P.O Box 230-80108, Kilifi, Kenya; School of Public Health, University of the Witwatersrand, 27 St Andrews Road, Parktown 2193, South Africa; MRC/Wits Rural Public Health and Health Transitions Research Unit (Agincourt), University of the Witwatersrand, Faculty of Health Sciences, Johannesburg, South Africa; Department of Psychiatry, University of Oxford, Warneford Hospital, Warneford Ln, Oxford OX3 7JX, United Kingdom; Department of Public Health, Pwani University, P.O. BOX 195-80108, Kilifi, Kenya; Institute for Human Development, Aga Khan University, P.O. BOX 30270-00100, Nairobi, Kenya

## Abstract

**Background:** Kenya, like many sub-Saharan African countries (SSA), is experiencing a rise in the number of HIV infected adults aged ≥50 years (recognized as older adults living with HIV [OALWH]). This trend has created a subgroup of vulnerable older adults demanding a prompt response in research, policy, and practice to address their complex and transitioning needs. Unfortunately, little is known about the health and wellbeing of these adults in Kenya. As such, we explore the experiences of OALWH and key stakeholders at the coast of Kenya to understand the health challenges facing the OALWH.

**Material and methods:** We utilized the biopsychosocial model to explore views from 34 OALWH and 22 stakeholders (11 health care providers and 11 primary caregivers) on the physical, mental, and psychosocial health challenges of ageing with HIV in Kilifi County, Kenya, between October and December 2019. Data were drawn from semi-structured in-depth interviews, which were audio-recorded and transcribed. A framework approach was used to synthesize the data.

**Results:** Symptoms of common mental disorders (e.g. stress, worry, thinking too much), comorbidities (especially ulcers/hyperacidity, hypertension, visual and memory difficulties), somatic symptoms (especially pain/body aches, fatigue, and sleep problems), financial difficulties, stigma, and discrimination were viewed as common across the participants. Suicidal ideation and substance use problems (especially ‘*mnazi*’ – the local palm wine and ‘ugoro’ – snuff) were also raised. There was an overlap of perceived risk factors across the three health domains, such as family conflicts, poverty, lack of social support, stigma, and the presence of comorbid health complaints.

**Conclusion:** Our findings provide a preliminary understanding of challenges, using the biopsychosocial model, facing OALWH in a low-literacy Kenyan setting. We found that OALWH at the Kenyan coast are at risk of multiple physical, mental, and psychosocial challenges, likely affecting their HIV treatment and overall health. Before programmes can have any lasting impact on these adults, improved access to basic needs, including food, financial support, and caregiving, and a reduction of stigma and discrimination must be addressed. Future research should quantify the burden of these challenges and examine the resources available to these adults before piloting and testing feasible interventions.

## Introduction

The last decade has witnessed a dramatic shift in the demographic profile of people living with human immunodeficiency virus (HIV) globally. Presently, many HIV clinics are caring for a growing number of adults aged ≥50 years (categorised as older adults) due to increased survival in people living with HIV (PLWH) and a steady rise in HIV diagnoses in this age cohort (1). In High-Income Countries (HICs), more than 30% of the PLWH are now aged ≥50 years (1), and this proportion is projected to reach 70% by 2035 in certain areas, including the United States of America (USA) (2) and some parts of Europe (2, 3). Among PLWH in sub-Saharan Africa (SSA), 15% are aged ≥50 years, and this proportion is expected to double by 2040 (4). In East Africa, Kenya has an estimated HIV prevalence of 9% (for those aged 50-54 years), 8% (among those 55-59 years), and 6% (for those aged 60-64 years), compared to the national HIV prevalence of 5% (5). These statistics herald a new era in the HIV epidemic response, where the needs and demands of the OALWH can no longer be ignored, especially in Eastern and Southern Africa, home to more than half the number of OALWH globally (1). Existing evidence suggests that OALWH faces a unique set of health challenges, which may differ from those of young PLWH. In addition to physical health problems, these adults experience mental health burdens and challenges to their social wellbeing that impair their overall health (6, 7). They are especially at risk of geriatric conditions like increased frailty and reduced functional ability, which may hamper their ongoing engagement in care and treatment (6). Unfortunately, the population of OALWH continues to be underserved and marginalized in research, policy, and clinical practice, especially in SSA.

Research findings, mainly from HICs, indicate that OALWH present with an average of three comorbid conditions in addition to HIV, including medical diseases (e.g., diabetes, hypertension), mental health problems (e.g., depression, anxiety, substance use) and social challenges (e.g., stigma, loneliness, and lack of social support) (6-9). Depression is exceptionally high among OALWH, up to five times greater than in HIV uninfected older adults (10). Anxiety, which is commonly comorbid with depression, is also said to be prevalent in this population, as high as 65%, in addition to elevated levels of current and past use of alcohol and other substances (11). Moreover, over 50% of these adults also display signs of cognitive problems, which are commonly related to other comorbidities and behavioral risk factors (6). The observed mental health challenges reduce the quality of life of these adults and have important health implications. For instance, depression and substance use remain strong predictors of poor HIV treatment outcomes and suicidal ideation (11). Untreated comorbidities commonly related to ageing, such as hypertension, may also exacerbate cognitive impairments (12). Furthermore, as PLWH grow old, the interaction of HIV, age and diabetes may elevate cognitive decline and reduce quality of life (13). The physical health problems faced by OALWH may also be complicated by environmental and psychosocial challenges such as poverty, food insecurity and lack of support (11). Though we lack a good understanding of the etiology of the physical, mental, and social impairments among OALWH, impairments appear synergistic and are driven by multiple factors, including HIV and its treatment, the ageing process, and diverse psychosocial and structural conditions in this population (6).

Despite the evidence of complex health challenges related to ageing and HIV, little research has qualitatively examined how OALWH understand their health and care needs. To date, most studies of ageing and HIV in SSA are cross-sectional studies focusing on biomedical processes and outcomes and rarely provide local insight into the health and wellbeing of these adults (14). Qualitative studies are needed to better understand the experiences and needs of this diverse population, especially among low-literacy populations in low resource settings. This is especially important as many cohorts of OALWH are emerging for the first time across the SSA region, and the apparent variability in findings among existing studies, e.g. in the prevalence and determinants of chronic comorbidities (14). Apart from complementing quantitative studies in accurately documenting the burden and determinants of the health challenges in these adults, qualitative studies will shed light on the contextual factors to guide the development or adaptation and subsequent implementation of culturally appropriate interventions in this population. Overall, the few qualitative studies among OALWH in SSA are mainly concentrated in Uganda (15-19) and South Africa (20-24). Others are from Kenya, Eswatini and Malawi (25-28). In Uganda, ageing with HIV is seen as a daily challenge financially and socially (15-19). The key barriers to successful ageing with HIV in this setting include stigma, food insecurity, unmet healthcare needs, particularly for associated comorbidities such as common mental disorders. In South Africa, the crucial barriers to living with HIV at old age include food insecurity, unemployment, the double burden of stigma, access to transportation and healthcare (20-24).

In Kenya, qualitative research related to older adults has mainly explored the impact of HIV on older adults as caregivers. Emerging data suggest that OALWH in the country face complex challenges when seeking care, including visits to multiple providers to manage HIV and comorbidities, poor services in HIV clinics, ageist discrimination, inadequate social support, and poor patient-provider communication (26, 27). However, these data come from the Western region of Kenya. As such, the health and wellbeing experiences of OALWH from other parts of the country with different cultural and ethnic groups, e.g. the Kenyan coast, is not known. Nevertheless, anecdotal reports from HIV clinics in this setting indicate a rising number of OALWH and several unmet healthcare needs. To bridge this research gap, we conducted in-depth interviews to explore the health challenges faced by OALWH at Kilifi, a low literacy setting at the coast of Kenya. Using the biopsychosocial framework, we explore the perceptions of 34 OALWH and 22 local stakeholders (11 healthcare providers and 11 primary caregivers) on the physical, mental, and psychosocial challenges of ageing with HIV in this setting.

### Theoretical Framework

We utilized Engel’s biopsychosocial (BPS) model of health (29), which provides a logical account of the chronic, complex, and dynamic nature of HIV. The fundamental tenet of this model for OALWH is the recognition of healthy ageing as the ability to thrive in an evolving environment influenced by physical/biological, mental/psychological, and social factors (as shown in Figure 1). The model provides a holistic approach to understanding the health needs of older adults and is supported by calls for research that positively impact the physical, mental, and social aspects of ageing with HIV (30-32). This model conceptualizes the physical, psychological, and social (both interpersonal and contextual) effects on health as dynamics. According to this model, the cause, manifestation and outcome of wellness and disease are determined by a dynamic interaction between physical, psychological, and social factors (29). Each component of the model includes systems that reciprocally influence other dynamics in the model and also affect health.

**Figure 1.** Components of the Biopsychosocial model of health

## Methods

### Study context and participants

This study was carried out between October and December 2019 at the Kenya Medical Research Institute-Wellcome Trust Research Programme (KWTRP) located within Kilifi County at the coast of Kenya. By the end of 2019, there were roughly 1.5 million residents in Kilifi County, the majority of whom were rural dwellers (about 60%), and 11% were aged ≥50 years (33). Most Kilifi residents belong to the Mijikenda ethnic group, whose main form of livelihood is subsistence farming and small-scale trading. Close to 60% of Kilifi County residents live below the poverty line, and one-third (36%) have not attained any formal education (34). Kilifi has an HIV prevalence of 38 per 1000 people (35). Women are disproportionately more affected than men (HIV prevalence of 54 vs 23 per 1000, respectively). The HIV prevalence for older people ≥50 years is unknown in this setting as it is not routinely documented during surveys. However, anecdotal reports from several HIV clinics in the study setting suggest an increasing number of older people living with HIV with unmet healthcare needs. Kilifi County is primarily served by one major public referral hospital (the Kilifi County Hospital), a few sub-county public hospitals and health centres. People living with HIV usually receive care in specialized HIV clinics within the primary facilities.

This study involved the following three groups of participants:

a. HIV infected older adults aged at least 50 years receiving HIV care and treatment at the HIV Comprehensive Care Clinic of the Kilifi County Hospital (KCH).
b. Healthcare providers attending to the older adults living with HIV. These included clinical officers, nursing officers, counsellors (within primary care facilities) and project managers of community-based organizations.
c. Primary caregivers of the older adults living with HIV who mostly tended to be close relatives, including spouses, children, and extended family members.

### Recruitment and eligibility

Study participants were selected purposively to represent diversity in the participants’ characteristics, including age, sex, residence, and the cadre of service (for healthcare providers). Initial contact was made with a community health volunteer (stationed at the Kilifi County Referral Hospital) with over ten years’ experience working with PLWH at the community and hospital levels. We capitalized on her experience and connections with several HIV clinics and community-based organizations around Kilifi during our recruitment process. Recruitment was conducted by a trained research assistant in liaison with the community health volunteer during their routine clinic visits. Participants expressing interest were then given more information concerning the study and how to take part. OALWH had to be ≥50 years old and on HIV treatment to be eligible. Primary caregivers were identified through the older adults living with HIV, usually during their scheduled clinic visits. These caregivers had to be directly involved in providing care and support, e.g. medical care to an older adult living with and spending time with the adult. HIV providers were approached at their place of work and invited to participate. We specifically targeted providers who provided direct care to OALWH. Participants who agreed to participate were then invited to a face-to-face interview at a convenient place, usually at KEMRI Kilifi.

### Data collection and tools

The lead author (PNM) conducted in-depth interviews lasting on average 45-60 minutes with each participant. All the interviews were guided by a pre-tested semi-structured interview schedule developed with guidance from prior work on HIV and ageing. All interviews were conducted in Swahili, English or Giryama. We also sought permission from the participants to take notes and audio record the interviews. A total of 56 interviews were conducted (34 among older adults living with HIV, 11 among healthcare providers and 11 among primary caregivers).

Among the older adults living with HIV, topic guide themes included: patient illness experiences following diagnoses, e.g. significant mental trauma, health changes over the years and the prevailing health challenges. These issues were explored using general open-ended questions, followed by additional probing to highlight further the issue raised. For healthcare providers, participants were invited to share their experiences providing care to older adults living with HIV. Specific topic guide themes included the important health challenges observed in the older adults and their preparedness to meet their needs. Among caregivers, we explored different issues, including their experiences taking care of OALWH and the challenges these adults face in their environment. We also collected respondents’ demographic information such as age, sex, educational status, and employment status. For HIV infected adults, we also collected HIV-related information, including years living with HIV. All participants were given Ksh 350 (about US$3) as compensation for time spent in the research, and transport expenses were also reimbursed.

### Data analysis

All the audio-recorded interviews were transcribed verbatim by a team of four trained research assistants. Subsequent data management was done using NVivo software (version 11). We applied the framework approach to analyze our qualitative data as described by Ritchie and Spice (36). Initially, two authors (PNM and AA) developed a preliminary coding framework inductively through in-depth reading of transcripts and deductively by considering themes from the interview schedules. These codes were discussed, and consensus reached on how they should be brought together into themes with guidance from the biopsychosocial model. The initial coding framework was progressively expanded to capture emergent themes as coding continued. When the coding was complete in NVivo, the lead author grouped all themes related to a specific concept together to form categories and exported this to a word-text processor to produce charts. The generated charts were used to summarize data, look for similarities or differences and explore patterns among the three groups of participants in the analyzed data.

### Ethical considerations

Written informed consent was obtained from all participants in the study. The study also obtained ethical clearance from the Kenya Medical Research Institute Scientific and Ethics Review Unit (KEMRI/SERU/CGMR-C/152/3804) and the Kilifi County Department of Health Services (HP/KCHS/VOL.X/171).

## Results

### Sample characteristics

We interviewed a total of 56 participants in this study (34 OALWH, 11 healthcare providers and 11 primary caregivers). Among OALWH, 53% were women; most (82%) had up to a primary level of education, most (73%) lived in multigenerational households, and their age ranged from 50 – 72 years. In addition, all the OALWH were on HIV treatment and had lived with HIV for a median duration of 12 years [10 – 15 years]. Healthcare providers included six registered nurses, two clinical officers, two project managers of community-based organizations and one HIV counsellor. All the primary caregivers were family members, the majority (73%) of whom were female. Further sociodemographic information and HIV-related characteristics of the OALWH are summarized in Table 1.

**Table 1.** Sociodemographic and clinical characteristics of older adults living with HIV

| Characteristic | n (%) or median (IQR) |
| --- | --- |
| Median age | 57 (54 – 63) |
| Female | 18 (53%) |
| **Educational level** |  |
| None | 7 (20%) |
| Primary Level | 21 (62%) |
| Secondary level | 5 (15%) |
| Tertiary level | 1 (3%) |
| **Type of employment** |  |
| Unemployed/not working | 12 (35%) |
| Small scale trader | 15 (44%) |
| Casual worker | 4 (12%) |
| Professional/skilled work | 3 (9%) |
| **Marital status** |  |
| Never married | 1 (3%) |
| Married | 19 (56%) |
| Separated/Divorced | 7 (21%) |
| Widowed | 7 (21%) |
| **Living arrangements** |  |
| Alone | 5 (15%) |
| Single generation household | 4 (12%) |
| Multigenerational household | 25 (73%) |
| Median household size | 5 (2 – 9) |
| Residence (rural) | 30 (88%) |
| Median duration since HIV diagnosis (years) | 12 (10 – 15) |
| On ART treatment | 34 (100%) |
| **HIV status disclosure** |  |
| Non-disclosure | 1 (3%) |
| Partial disclosure (to close family only) | 18 (53%) |
| Full disclosure (beyond close family) | 15 (44%) |

**Notes:** ART – Antiretroviral treatment; IQR – Interquartile Range

### Perceived biopsychosocial challenges confronted by OALWH

The different forms of biopsychosocial challenges faced by OALWH are summarized in Table 2. Overall, most of the OALWH shared their 10-plus years of experience living with HIV, from the time of diagnosis to their current state. The majority of the OALWH described moving from a state of shock, fear, anger, denial, hopelessness, or suicidal ideations upon diagnosis to a state of acceptance and ownership. This transition would not be without challenges, as many of the OALWH noted having gone through traumatic experiences from family members, friends, workmates, and healthcare providers. These traumatic experiences, e.g. pervasive stigma (both enacted and internalized), prolonged periods of illness, spousal abandonment and separation, and financial loss, led to chronic stress and low mood. Thereafter, they narrated moving from a state of ownership to a state of constant survival characterized by an array of health problems which we have conceptualized as physical (biological), mental (psychological) and social according to the biopsychosocial model. Most participants believed that older adults living with HIV in Kilifi face an intricate mix of physical, mental, and psychosocial challenges. These challenges are described in detail in the following sections.

**Table 2.** Perceived forms of physical, mental, and psychosocial health challenges facing older adults living with HIV as discussed by study participants at the coast of Kenya

| Forms of health challenge | Number of in-depth interviews (among OALWH) where the health challenge was discussed (N = 34)<br>n (%) | Number of in-depth interviews (among caregivers) where the health challenge was discussed (N = 11)<br>n (%) | Number of in-depth interviews (among healthcare providers) where the health challenge was discussed (N = 11)<br>n (%) |
| --- | --- | --- | --- |
| **Physical complaints** |  |  |  |
| Comorbidities: chiefly ulcers/hyperacidity, hypertension, and diabetes | 29 (85%) | 9 (82%) | 11 (100%) |
| Wide-ranging physical symptoms, e.g. pain/body aches, fatigue, insomnia | 33 (97%) | 10 (91%) | 9 (82%) |
| Functional impairments, e.g. difficulties walking, writing, holding things | 18 (53%) | 3 (27%) | 3 (27%) |
| **Mental complaints** |  |  |  |
| Symptoms suggestive of common mental disorders, e.g. thinking too much, worry, stress, low mood, hopelessness/giving up easily | 29 (85%) | 9 (82%) | 9 (82%) |
| Cognitive complaints chiefly memory difficulties. Others including slow mental processing/learning, attention/concentration problems | 22 (65%) | 7 (64%) | 4 (36%) |
| Current drug and substance use, mainly '*mnazi*' and '*ugoro*', which are the local palm wine and smokeless tobacco, respectively | 5 (15%) | 0 (0%) | 9 (82%) |
| Suicide and suicidal ideations | 3 (9%) | 0 (0%) | 4 (36%) |
| Psychotic like symptoms, e.g. disorganized behaviour, aggression, lack of restraint and unwanted thoughts. | 1 (3%) | 3 (27%) | 3 (27%) |
| **Psychosocial complaints** |  |  |  |
| Financial difficulties, e.g. lacking school fees for dependents, walking long distances for HIV care | 31 (91%) | 8 (73%) | 9 (82%) |
| HIV-related stigma and discrimination especially internalized and enacted stigma | **29 (85%)** | **8 (73%)** | **11 (100%)** |
| Ageism/ageist stereotype e.g. neglect, verbal insults, vandalism | **19 (56%)** | **5 (45%)** | **7 (64%)** |
| Isolation and lack of support | **19 (56%)** | **5 (45%)** | **8 (73%)** |
| Loneliness | **17 (50%)** | **1 (9%)** | **8 (73%)** |
| Food insecurity, e.g. skipping meals or going days without food | **16 (47%)** | **5 (45%)** | **9 (82%)** |

### Perceived physical health challenges

Physical complaints were described to be common among adults ageing with HIV by several participants. Key among these concerns was the onset of multiple comorbidities in these adults, which was frequently associated with pain, loss of control of one’s body, emotional distress, and increased treatment burden (e.g. attending multiple clinics and medication burden). Hypertension, diabetes, ulcers/hyperacidity, hearing, and visual impairments were the most frequently reported comorbidities across the participants. Other conditions discussed (though to a lesser extent) included obesity, arthritis, stroke, teeth problems, cervical cancer, TB, and pneumonia. Apart from these comorbidities, somatic symptoms were also discussed in several interviews across the groups. Pain (in the back, extremities, joints, chest, whole-body), headaches, insomnia, fatigue or low energy, and numbness in the limbs were frequently reported. Sometimes, these symptoms were associated with emotional distress and functional impairments, e.g. difficulties bathing, using the phone, and performing house chores. Some participants believed that the onset of the physical challenges was because of HIV-related factors (e.g. HIV infection itself, long-term ART), multimorbidity, old age, food insecurity, emotional distress, substance use, and the nature of someone’s work. Participant quotes are given in Table 3.

**Table 3.** Participants’ physical health challenges quotes.

| Perceived health problem | Examples |
| --- | --- |
| Multiple comorbidities | I am weak and constantly fatigued because I have ulcers, high blood pressure in addition to diabetes. My eyes are also painful and feel strained, and I sometimes experience toothache. As we are talking right now, my ears are not hearing well. So, all these diseases make me weak and significantly affect my well-being (**OALWH**) |
|  | Ulcers is their main problem. Many of them complain of ulcers, and this is mainly because of stress. Anything small affects them. Sometimes you find one thinking too much, and another telling you, 'my children do not take care of me, they have forgotten about me', or 'I have one child.' Someone tells you about their child who died a long time ago but is still affected. Stress is a big problem because they pile up issues, not because of HIV but the other issues. Another issue is food. The elderly must look for food for themselves and their irresponsible children, who may be drunkards. Often, you find that they deal with their stress and that of their children (**Healthcare Provider**) |
|  | Last year, my hands were painful, and I could not bend. I carried out an X-ray and was told it was arthritis. I was told there was no treatment, and the only relief was by massaging myself and using painkillers. So, I bought clove oil, and it is what I have been using. There are improvements, but the problem is not resolved yet. I am also unable to work and to make it worse, I have difficulties sleeping whenever I am stressed. I was recently diagnosed with high blood pressure but am not taking medications yet. I also have ulcers, but I do not usually keep it in mind (**OALWH**) |
| Somatic symptoms | My knees are painful. Sometimes even squatting is a big challenge. I also cannot rise too quickly from a seated position; I have to support myself. I must also use painkillers for some relief before I can work. This has affected me because I cannot work without them. Whenever the analgesic effect wears out, it becomes very difficult to use my legs. I also experience headaches sometimes, and my relief is usually the painkiller. These pain killers have become my life now (**OALWH**) |
|  | My pace has become very slow nowadays. I feel heavy, and there is nothing I am lifting or carrying. I often feel tired, and, in such times, I prefer walking around for some time. I must do this to ward off intrusive thoughts. Since I became diagnosed, I have also grown weak and have lost my libido. In fact, whenever I start having sex, I have this sensation of my waist being cut with a razor (**OALWH**) |
|  | My spouse cannot do heavy manual jobs. He used to work at the sea at night, which is usually cold and requires energy. It reached a point I had to stop him from going there anymore because he was weak and felt he was risking his life. Before this, he often used to complain of being sick. I was not at peace. I found it pointless for him to earn money that way and end up using it to medicate him. So I told him to stay at home and do anything else. We are living by the mercies of God (**Caregiver**) |

### Perceived mental health challenges

Participants discussed various mental health problems facing OALWH in the study setting. We have discussed these issues under the following headings: symptoms of common mental disorders, cognitive symptoms, substance use problems, and others (suicidal ideation and psychotic symptoms). Notably, symptoms of common mental disorders were more frequently discussed than the other problems. Participant quotes are given in Table 4.

**Table 4.**
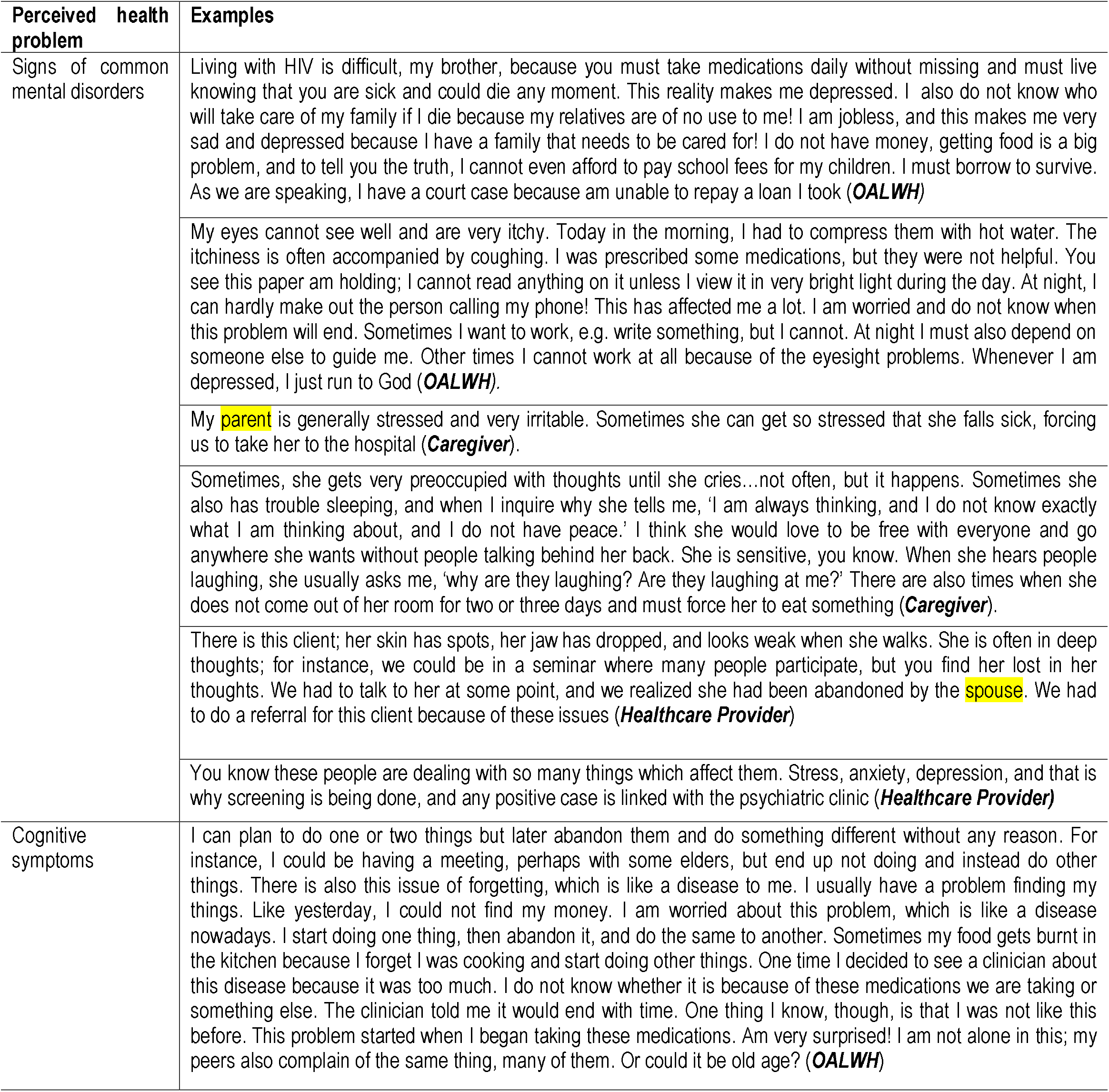
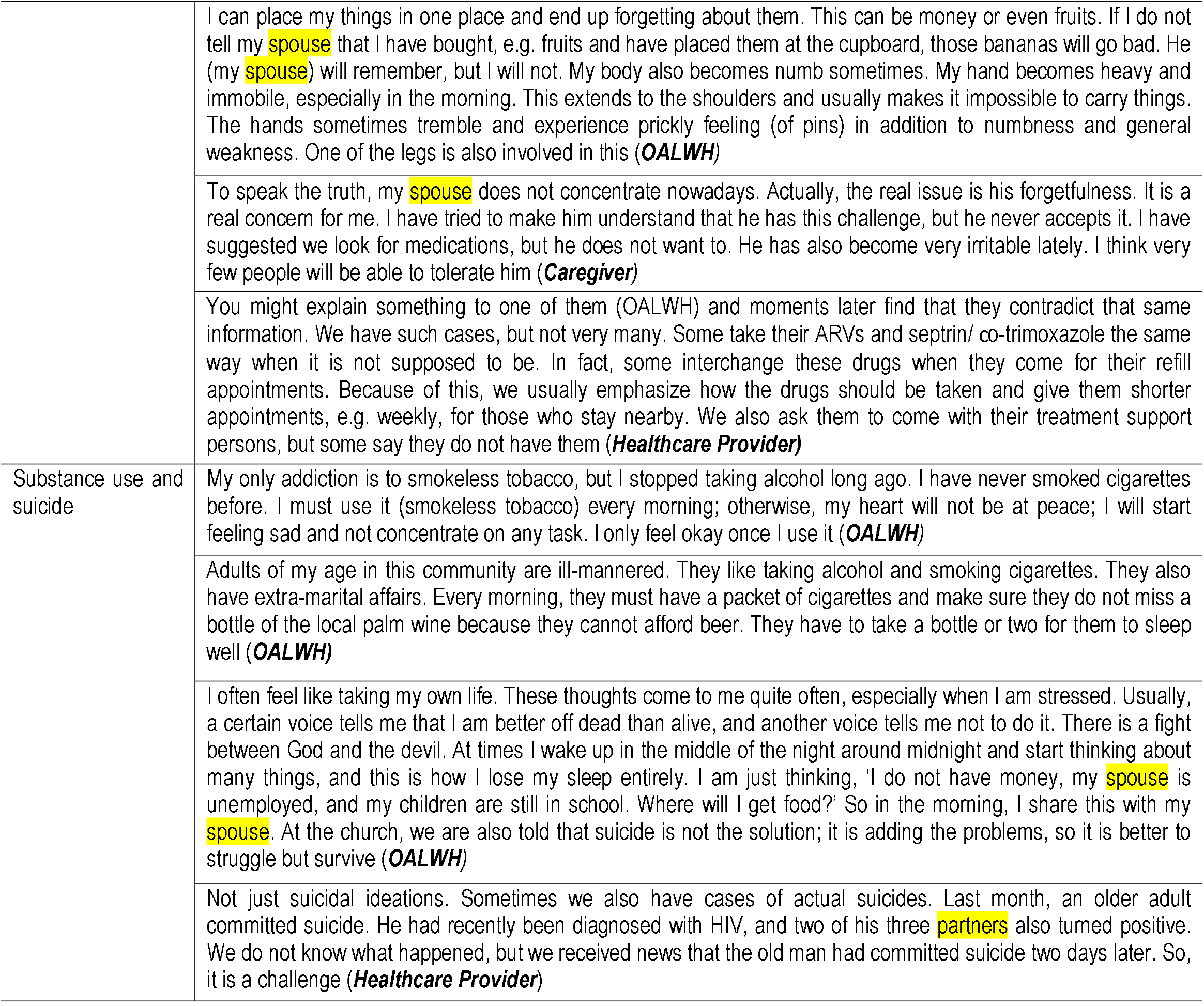
Mental health quotes

#### Symptoms of common mental disorders

Emotional and behavioral symptoms, suggestive of anxiety and depression, were described to be prevalent across the participant groups. Key among these symptoms included thinking too much (especially at night or when alone), persistent stress, low mood, anger/irritability (for no apparent reason), worrying a lot, hopelessness, restlessness, panic attacks, and nightmares.

Other prominent symptoms included insomnia (difficulty falling asleep or interrupted sleep), self-isolation, low self-esteem, headaches, and low libido. Overall, these symptoms were described to be on-and-off among the OALWH and were frequently associated with the exacerbation of physical conditions (such as hypertension), chronic use of drugs (especially analgesics and sleeping pills), financial instability and overall poor quality of life. For many participants, the onset and persistence of physical body changes (e.g. peripheral neuropathy, obesity, body weakness, fatigue, pain, vision, and hearing difficulties) were considered a significant risk for the mental complaints. A few OALWH also associated their mental symptoms with evil spirits. A limited number of OALWH sought medical attention for the highlighted issues. They considered these symptoms a ‘normal way of life’ while many others did not know what to do or lacked the means to access any help.

#### Cognitive/neurological challenges

The commonly reported complaint in this category was memory difficulty, e.g. difficulties recalling important dates or finding items (money, keys, foodstuffs). Surprisingly, most OALWH reported that they rarely forgot to take their medications as the practice had become part of their lives. Many healthcare providers corroborated this, although they also noted that there are a few OALWH at risk of poor adherence caused by cognitive impairments. For instance, some OALWH forgot to take their medications (especially those taking mnazi) while others mixed up their medications (e.g. ARV drugs and co-trimoxazole). Other cognitive symptoms included attention/concentration, processing/learning, movement, and multi-tasking difficulties. Peripheral neuropathy (manifesting as numbness or pain in the extremities and difficulty holding items) was also reported to be a common neurological problem in these adults and was associated with significant distress. Among OALWH, the cognitive symptoms were viewed as an additional disease, and many did not understand its causes. However, they bemoaned the considerable impact it had on their lives, such as pain, reduced efficiency in doing daily responsibilities (e.g. reading, bathing, eating), worry and anxiety, decreased productivity and caregiver distress. The few who sought medical attention ended up being disappointed, sometimes advised that it will resolve on its own, or there is medicine, but it is costly.

#### Drugs and substance use problems

Current drug and substance use was mainly discussed by healthcare providers. In contrast, most OALWH (especially men) reported having a history of drug and substance use, especially alcohol and tobacco but stopped following HIV diagnosis. However, some confessed to being social drinkers. ‘*Mnazi*’ (the local palm wine and ‘*ugoro’* (smokeless tobacco or snuff) were the commonly used substances. Many providers emphasized that *mnazi* drinking is a big problem (both for men and women), noting that some OALWH came for their routine clinic appointments while drunk and sometimes forgot to take their ART drugs. Some providers also associated the *mnazi* drinking problem with unsuppressed viral loads, treatment default, sexual risk-taking behaviors, e.g. multiple partners, cross-generational sex (especially in the mnazi drinking dens dubbed ‘*mangwe*’). The key underlying factors for the increased use of *mnazi* included family conflicts which often led to stress, unstable sexual partners, negative coping skills, culture, working in a mangwe and the easy access of these substances (cheap and readily available). *Ugoro*, the other commonly abused substance, was especially common among women. It emerged that some of the *ugoro* users had become dependent on the substance. Beer, cigarettes, khat and bhang were also discussed, although to a lesser extent.

#### Suicidal ideation and psychotic-like symptoms

Three OALWH reported having suicidal ideations, which was intermittent. This was corroborated by the healthcare providers. The commonly associated factors included persistent stress (caused mainly by financial difficulties and family conflicts), evil spirits, HIV-related (e.g. getting tired of taking ART and not getting healed). One healthcare provider reported a recent case of suicide involving one polygamous man who had been diagnosed with HIV together with two of his three wives. Though limited, there were also few reports of psychotic-like symptoms (e.g. disorganized behaviour, aggression, lack of restraint and unwanted thoughts) across the three groups of participants. These cases were frequently observed around the time of HIV diagnosis. After that, the symptoms subsided, but the OALWH continued taking antipsychotics. Three participants were reported to be on these medications in the interviews.

### Perceived psychosocial challenges

Participants discussed a range of psychosocial challenges. We present these issues under the following categories: a) financial challenges, b) stigma and discrimination, c) loneliness and isolation. Participant quotes are given in Table 5.

**Table 5.** Participants’ psychosocial challenges quotes.

| Perceived health problem | Examples |
| --- | --- |
| Financial challenges | I am jobless, my brother. I have also lost my hands-on skills and experience with time. I am weak and do not have money. Getting food is a problem, and to speak the truth, even raising school fees for my children is a huge problem. Nevertheless, I have a very good friend, who is of great help to me. Sometimes, I eat at his place, lunch, and dinner, but he does not know I live with HIV. Sometimes I am depressed and anxious because I do not know how to provide for my family. My relatives are of no help to me. Like yesterday, we did not have dinner, and I had to borrow the fare to come to this place. As we speak right now, I am in debt and face a court case because I cannot repay a loan. Often, I sleep hungry and must borrow to survive (*OALWH*). |
|  | To speak the truth, I do not receive any support from my relatives, and this is not because there are no people to help. Many of my relatives have died. I often sleep hungry. Sometimes, I only have a cup of porridge for three consecutive days...and this too is usually borrowed from a neighbor so that I can take my medications. Other times, I survive on a cup of tea or hot water. Fare is also a big challenge. However, whenever I lack fare, I usually walk to the clinic (around 50 km journey). Oftentimes, I start my journey two days ahead of my clinic appointment and have two rest points in between (*OALWH*) |
|  | Financial challenge is a big problem for these adults because they are neglected by their children sometimes.<br>The day I am on duty at the clinic, I usually give many of them fare. You will hear many saying, 'sister, please help me with fare, I walked to the clinic, and I have to walk back.' If you have a good look at them, they appear exhausted, so you end up helping them. Even food is a challenge. In our discussions, the majority say that they have one meal per day and mind you, some fail to take their medications whenever they miss a meal due to dizziness claims (*Healthcare Provider*) |
|  | The other day, we visited an elderly client in the community, taking care of two children living with HIV. She is unemployed, and the children's **parent** is not around. The children are in school. They need support with their schooling in addition to food. She is largely unable to provide all these and often must borrow. Sometimes, you find that even the skills you are giving them, e.g. rearing chicken or fish, and making baskets, are not helpful because they are old and unable to work. In such cases, we assist them with food, counselling sessions, linkages, and referrals to foster treatment adherence and the well-being of the children **(Healthcare Provider)**<br>I usually borrow money from neighbours or friends to buy food whenever my **parent** does not have. I do not mind the debt because my target is to ensure that my **parent** has food to take her medications. However, it is a bit stressful because sometimes she sleeps hungry. And this is complicated by my other **parent**, who often quarrels her with small issues, and this increases her stress. Every time my she asks for money, he says he does not have it, and if we ask him how will we survive, he replies, 'can't your child look for the money?' It is frustrating because how can I take up this responsibility when I am still in school? In fact, the one educating me is usually **another distant relative (Caregiver)** |
| Stigma and discrimination | The elderly living with HIV undergo many challenges in the community. For instance, you find one is sick, and no one is taking care of him at home. In fact, people will avoid interacting with this person, especially where he sleeps, out of fear of contracting the virus. Some of them have called me seeking help. Sadly, when I go to their homes, I find some have not even taken a bath for a whole week! In such a case, I usually clean up the person and assist him to eat. It is sad because the family just places the food there; the person is sick and can hardly eat by himself! Some are told to stay with their HIV because they know where they took it from! As we are speaking, there is a woman I would like to talk to because she stays at XX place but takes her ART medications from a faraway place because she does not want relatives to know that she is living with HIV. So, HIV education has come for sure, but it is far from being complete **(Healthcare Provider)**<br>She is often in low moods. Whenever she hears people talking and laughing on the other side, she gets offended. I think she would really like to be free and move about without people badmouthing her. **(Caregiver)**<br>For men, there is a challenge, a huge challenge. Getting them is a very big challenge. We have tried to work with male champions to convince their fellow men to join us. Stigma is the big issue. In our meetings, you mainly find the ladies coming, and when you ask them the whereabouts of their **spouses**, they often tell you, 'He has gone to work and has sent me to listen to the messages on his behalf.' So, they have that fear; they do not want to be seen. The case is not different in the primary facilities because many of them are not coming. He either sends the **partner** or comes but is very careful not to be seen, e.g. comes in through the back. Because of this fear, you find that some of the men do not want to be tested. We have several cases where **one partner** is HIV infected; the one got tested and turned positive but is still in denial. You will find one of them saying, 'we went and got tested with him but has insisted that the machines were not functioning and as such he is not infected.' Others also note that their **partners** have constantly refused to start ART and use condoms, and whenever these issues are brought up, they end up quarrelling **(Healthcare Provider)**<br>That is a big problem (stigma). Ours is a small facility, so we schedule one day in a week to see the clients living with HIV. Surprisingly, even the community knows that Tuesday is the day for those taking ARV drugs. So, most of them come very early in the morning. Therefore, I must come in very early to serve those who have stigma. Alternatively, that person will hang around the clinic from morning till evening when everyone has left so that people will not understand what she is doing there, or you will find that they will skip their appointed clinic dates **(Healthcare Provider)** |
|  | Some of them are treated very badly when they come for their appointments until I get angry myself! Let's say you are late for your appointment; you will be shouted at, insulted, and reprimanded like a child. In fact, some end up crying and I have to calm them down. It is sad because the culprits are usually the **healthcare providers** who often shout at them, saying, 'Take your drugs, or you will die of AIDS alone, and you won't see us!' It is shocking because some of them are treated very badly to the point that I am also angered. Some of them come to my desk shedding tears, so I have to calm them down. At some point, it was too much that some of them transferred to other clinics (**OALWH**)<br>The elderly are being burned alive and cut with pangas. As a matter of fact, some of my neighbours recently cut their **elderly relative** severally with a panga because of witchcraft accusations. Later, **another relative** also passed through the same ordeal...the head was completely slashed off with a very sharp panga! And the reason is always witchcraft; I have not heard of any other thing. Such cases are numerous where I am living. I have witnessed four elders being cut with pangas, and the sad part is that these people will never expose each other to the police...even if I know it is you who did it, I will never say it. Even when the police come, the locals will never expose the culprit (**OALWH**) |
| Loneliness and isolation | I really desire to have an exclusive male companion of my age to spend the rest of my life with. Having one who is already married is problematic because sometimes I am very lonely and would want to call him, but I end up avoiding it out of fear, 'what if he is with the **spouse**? And you know it is not good to break someone's marriage.' Because of such challenges, I would like to have my own man who does not belong to anyone, one I can relate to freely and love fully. You know, with someone else's **partner**, the love is incomplete, and it feels like I must hide constantly, and it reaches a point I feel afraid. Sometimes he does not pick up my calls and sends a text message saying he is at home. So, you see, with such problems, I just want to get my own man who will live with me, someone I can talk with, especially when things are difficult because it is stressful at times (**OALWH**)<br>Sometimes, I wish I could have friends with whom I can confide and share my challenges, but I am afraid of being discriminated. You cannot trust anyone. I have many friends, but they are all fake (**OALWH**)<br>There are times he isolates herself from other people. You might see her seated alone and in deep thoughts. Sometimes she wakes up and begins to shed tears. She usually feels that her life has come to an end, perhaps (**Caregiver**)<br>Whom will I share my problems with? I do not have anyone. Anyone I open myself to will want to know how and why my **spouse** is facing those challenges. What is the cause? This will mean I start discussing and revealing the issues of my **spouse**, which is not good (**Caregiver**) |

#### Financial challenges

This was discussed in virtually all the interviews we conducted. Many participants felt that financial difficulty was the most crucial issue affecting OALWH in the study setting because of its massive impact on food security, caregiving responsibilities, emotional well-being, and management of HIV and other comorbidities. Many OALWH reported skipping meals or going hungry for some days (sometimes up to three consecutive days), lacking school fees for their dependents, and walking long distances to access HIV care (sometimes 50 kilometres) for lack of money. This was corroborated by the healthcare providers and caregivers, who further stated that many of those who slept hungry tended to skip their medications, complaining of dizziness, headaches, and stomach discomfort. To some extent, this was associated with poor treatment outcomes, including unsuppressed viral load and poor retention in care. Several factors were perceived to bring about these financial challenges, including neglect/lack of support, reduced productivity (e.g. because of physical weakness), caregiving responsibilities, cost of managing HIV and other comorbidities (e.g. attending multiple clinics) and unemployment. To make ends meet, a good number of the OALWH resorted to borrowing money from friends or taking small loans (e.g. mobile loans), which sometimes they failed to repay, leading to conflicts.

#### Stigma and discrimination

This theme incorporates HIV-related stigma and ageism (discrimination based on age). Despite relatively high levels of disclosure, a significant number of OALWH experienced HIV-related stigma. Discriminatory behaviour (enacted stigma from malicious gossip to outright discrimination, e.g. neglect, isolation, verbal insults) were reported to be common, especially in the most rural areas of the study setting. The perpetrators were mainly family members. Stigma was also reported to be common in mnazi drinking dens ‘mangwe’, burial ceremonies, HIV clinics and certain churches. Internalized stigma (e.g. feelings of shame, guilt) emerged as an important theme, especially among the OALWH and was frequently associated with self-isolation, anxiety or high consciousness of self, fear of seeking assistance, attending very far HIV clinics, stress, and irritability. Many participants also noted that a few community members hold negative perceptions (relatively passive but leading to acts or habits such as gossip) about OALWH, which seemed to compound the internalized stigma among OALWH. Overall, stigma was associated with physical deterioration, persistent stress, non-disclosure, reduced access to support services, and poor treatment outcomes.

A significant number of the participants also highlighted discrimination based on age, which was perceived in multiple settings, including the home (mainly through isolation, neglect, lack of respect from children), HIV clinic (e.g. scolded openly and verbal insults), and the community level. It also emerged that some of the community members around the study setting (especially in the most rural areas) regarded older people suspiciously. Many participants narrated hearing or witnessing several older people being beaten, beheaded, or burned alive in their houses for suspected witchcraft, and the perpetrators were mainly close family members.

#### Loneliness and isolation

This emerged as an important theme in the conversations with OALWH and healthcare providers. Despite living in multigenerational households, a significant number of the OALWH expressed loneliness and isolation. Some had lost their partners, some their closest relatives and others saw their circle of friends getting smaller. Still, others felt neglected by those around them. Some decided to stick to themselves out of fear of how the people would react if they sought help, as this would mean disclosing their status. As a result, such individuals described this situation as affecting their mental health with persistent sadness and stress.

## Discussion

### Summary of key findings

We conducted this study to gain a preliminary understanding of the health challenges facing OALWH at the coast of Kenya. Our study provides insight into the complex challenges of ageing with HIV in the study setting. It also opens opportunities for further epidemiologic research and subsequent development of tailored interventions for this vulnerable yet understudied population. Overall, our findings reveal that OALWH in this setting are particularly vulnerable to mental health problems, especially symptoms of common mental disorders, which sometimes progress to severe forms such as severe depression and suicidal ideation. They are also at risk of physical challenges, including comorbidities (especially hypertension, ulcers, and diabetes) and somatic complaints such as pain, fatigue, and insomnia. The mental and physical impairments are complicated by psychosocial challenges, including poverty, lack of support, stigma, and discrimination. Noteworthily, there was an overlap of perceived risk factors across the three health domains (e.g. family conflicts, poverty, food insecurity), suggesting that OALWH who experience these shared cumulative risk factors are more likely to face multiple health challenges. It also implies that the action taken to mitigate any or some of the shared enabling factors is likely to have a preventive spillover effect across multiple health domains. Most of the views of OALWH on health challenges were corroborated by those from the providers and caregivers. However, a few disparities emerged on some of the perceived health challenges. Current drug and substance use, for instance, was reported mainly by healthcare providers, while cognitive and somatic complaints were discussed by OALWH. By and large, our results indicate that OALWH increasingly require care that extends beyond HIV treatment, hence the urgent need to reprioritize health systems to meet their complex needs.

### Physical health challenges

Our discussions clearly showed that physical challenges are important concerns for older adults living with HIV at the coast of Kenya, given their negative impacts on other health domains and overall health. The onset of multiple conditions and somatic symptoms was especially associated with pain, distress, increased treatment burden, e.g. having to attend multiple clinics reduced quality of life and functional impairment. As the number of OALWH increases in many HIV clinics, HIV care will increasingly need to draw on a wide range of medical disciplines besides evidence-based screening and monitoring protocols to ensure the healthy ageing of OALWH (3). Unfortunately, the implications of such a trend could present a significant challenge for many SSA countries, including Kenya, where older adults’ care is fragmented. Most healthcare providers presently lack guidance and training to identify and manage declines in physical and mental capacities in this population. The siloed provision of HIV care and other comorbidities could also imply that providers are unaware of the patients’ other conditions. In the current study, many OALWH reported attending multiple clinics for different conditions, which constrained their already limited funds. Some OALWH had to choose between the ART clinic and the other clinic (e.g. hypertension and diabetes). However, some providers noted that they tried to streamline appointments among the outpatient clinics, but this was not always possible. Our findings are similar to those reported in a recent qualitative exploration of challenges with seeking HIV care services for OALWH (26). Specifically, OALWH raised unique challenges when seeking HIV care, including multiple visits to healthcare providers and poor patient-provider communication. Integration of services for HIV and non-communicable diseases in primary care may enable settings like Kenya to expand healthcare coverage for PLWH. Nonetheless, there is limited evidence of its effectiveness (37). Despite some challenges, e.g. inadequacies in structure, process and outcome, initial evidence shows that service integration can be a valuable strategy to improving health outcomes among PLWH (38).

The current findings also revealed a prominent intersection between ageing and HIV. For most participants, ageing rather than HIV was the primary concern. This is not surprising considering that among PLWH with controlled viraemia, HIV infection often stops being the overriding comorbidity but is simply a key element in the overall milieu of multiple conditions (39, 40). In a few instances, however, participants discussed that their experiences were associated with HIV, long-term medication use, or side effects, although their providers attributed these conditions to normal ageing. They advised the OALWH to ‘wait’, and they will improve with time. Disagreements about the cause of a symptom or health condition may contribute to doubts about the effectiveness of treatment or conceal an emerging disease and contribute to delayed diagnoses such as medication side effects and polypharmacy. A few of the OALWH in our study resorted to alternative forms of management, e.g. prayers and traditional medicine, after their conditions failed to improve following medical intervention. Overall, there is an urgent need to integrate other chronic diseases into HIV care and train staff on how best to attend to the unique needs of OALWH.

### Mental health challenges

Our findings of substantial reports of common mental disorder symptoms among OALWH are consistent with previous reports of poor mental health among PLWH in the study setting, albeit among younger populations (41, 42). However, our study does not establish whether the burden among OALWH is higher or lower than that observed among young PLWH. Quantitative studies are needed to confirm this comparison. Nonetheless, OALWH may be facing a higher burden of common mental disorders than their younger counterparts for different reasons. Firstly, many of the longest surviving OALWH may be significantly impacted by the legacy of the early years of the epidemic, including multiple bereavements, ‘survivor guilt’ and post-traumatic stress disorder. Living for many years with an uncertain prognosis, financial challenges, and a historical and ongoing stigma may indeed contribute to a high burden of anxiety and depression in this cohort (43). Secondly, the higher burden may also be attributed to the multiple challenges that OALWH face, e.g. poverty, food insecurity, caregiving responsibility, double-stigma, and the onset of physical body changes, as evidenced in our study. Our results are also partly corroborated by findings from other settings in SSA. For instance, Charlotte and colleagues reported a high prevalence of severe depression among OALWH in West Africa, especially among those who were unemployed, current or former smokers (44). In other studies, OALWH have been noted to have lower rates of common mental disorders than uninfected age peers or younger PLWH (45). The current study did not quantify these problems, and future studies need to do so.

Since the beginning of the HIV epidemic, the manifestations of cognitive and neurological problems have been ubiquitous and frequently associated with poor treatment outcomes and impairment of activities of daily living (11). In our study, the most frequently reported cognitive problem was memory difficulty, which was commonly associated with stress and shame. Strikingly, healthcare providers seldom suspected cognitive impairments among OALWH and screenings were never done. This is a huge blow in detecting and managing these symptoms and contributes to the increased burden of stress and discomfort in this patient cohort. This observation is similar to what has been reported in South Africa (40, 46). Despite the reported memory challenges in this study, OALWH rarely forgot to take their medications (from their own self-reports and that of their providers). This finding is not unique in the HIV literature (47). It is possible that OALWH are more organized and experienced, and possibly more motivated after experiencing the initial devastating outcomes of the HIV pandemic. There could also be a survivor effect, where those living longer are those likely to be more adherent while those that were more forgetful were non-adherent and succumbed to AIDS. However, it is still essential to monitor the cognitive function of these adults to prevent treatment non-adherence, considering the multiple challenges they face, which are likely to impact their cognitive function.

Our findings also noted a section of OALWH at risk of substance use dependence, especially home-brewed alcohol (for both men and women) and smokeless tobacco (mainly women). This is not surprising given that the consumption of mnazi (the local palm wine) is known to take precedence among the inhabitants of Kilifi because it is cheap and often less regulated (48). In the current study, OALWH residing in the most interior parts (away from periurban areas), those working in mnazi drinking dens (especially older women), and those experiencing family conflicts were regarded to be most at risk of this problem. While it is true that a significant number of OALWH stop taking drugs and substances following HIV diagnosis, as noted in previous studies (ref), there are still some who abuse drugs and substances, leading to poor overall health and treatment outcomes. The need to address this problem is even more crucial since it was associated with rising cases of sexual risk-taking behaviour in this cohort, including cross-generational sex, multiple sex partners and non-condom use.

### Psychosocial challenges

Psychosocial factors are well-known predictors of treatment adherence, disease progression and quality of life for PLWH. Stigma and loneliness, for instance, are highly linked with mental ill-health in OALWH (49). Unlike social determinants of health (e.g. economic stability and education), psychosocial factors are considered more malleable, resulting in improved health status (8). Hence, understanding the psychosocial needs of OALWH is critical to developing interventions aimed at improving their overall health. Findings from our exploratory study suggest that financial difficulties, loneliness, stigma, and discrimination are prevalent among OALWH in the study setting and are associated with mental complaints (e.g. persistent stress) and physical health problems (e.g. worsening other conditions and poor treatment outcomes). These findings are not surprising given the prevailing situation of older adults in the country, especially those residing in rural areas. According to Help Age Kenya, the majority of older people in Kenya, especially those in rural areas, live in absolute poverty (50). Besides, individuals aged ≥50 years are the poorest age group in the country, and the majority have no formal sources of income; hence no pension to fall back on in their old age. In Western Kenya, Kiplagat and colleagues identified lack of support, HIV-related stigma, ageism, poor patient-provider communication as some of the key challenges facing OALWH when seeking HIV care services (26). In our study, although many OALWH lived in multigenerational households, very few reported receiving instrumental support. A recent report, the National Gender and Equality Commission Report, dubbed ‘Whipping Wisdom’, also established that older adults in Kenya faced various forms of violence, including social stigma, neglect, abandonment, and hindrance from using and disposal of property (51). Similar to our study, the drivers of such violence include land succession disputes, accusations of witchcraft, poverty and degrading family and community values. Elder abuse is also recognized worldwide as a serious problem (52). The issue cannot be adequately resolved if the essential needs of older people (especially those at risk of poor health like OALWH) – for food, shelter, security and access to healthcare are not met. There is a need to create an environment where ageing is accepted as a natural part of the life cycle and where ageist stereotypes are discouraged.

### Implications

Our study highlights opportunities for interventions and further research. Most importantly, interventions for this unique population must be contextualized by both age and sex. It is important to reiterate that the health challenges faced by OALWH at the coast of Kenya are often interconnected and require a cohesive and collaborative response to achieve maximum benefits. Such interventions should target modifiable factors such as emotional support and integrate needed social and community support, e.g. case management services, food and nutrition support, financial assistance with caregiving responsibilities, and transportation. Context-specific interventions to help OALWH develop and nurture their own coping strategies are also critical in this population. Patient-centred care and patient self-management principles (e.g. self-reliance and empowerment) are critical elements in chronic care and are advocated as universal strategies in international frameworks of chronic care (53). While OALWH are the primary target of most of the existing interventions in this cohort (54-56), research is required on how to build the capacity of healthcare providers, family members who act as informal caregivers and friends to provide support and care to those ageing with HIV. Future work should also examine the impacts of poverty on HIV care for OALWH as our results indicate that financial challenges influence many of these problems. Yet, it is essential to note that despite the health challenges discussed by participants, all the OALWH were currently receiving HIV care. Future research should also examine the resources and resilience among these individuals to fully understand the vital role of resilience in empowering OALWH to enact processes that buffer health from the identified stressors. There is also a need for future research to quantify the existing burden of physical and mental challenges in this population in Kilifi and confirm the risk and protective factors to these challenges. Furthermore, there is an urgent need for research to pilot and test the applicability and effectiveness of interventions underlying the determinants of physical and mental impairments in this setting.

### Strengths and limitations

To our knowledge, this is the first study to richly explore the health challenges of OALWH at the coast of Kenya and among the few studies in Kenya. Unlike previous studies, a key strength of this work is that participants comprised a diverse group of stakeholders, including clinicians, community-based organizations, caregivers apart from the OALWH themselves. This ensured that the views were diverse and contrasted across the different participant groups. The use of a biopsychosocial model helped us to obtain a richer insight into the complexity of the identified health challenges across the three domains in Kilifi. However, our findings emanate from a predominantly rural setting, and circumstances may differ from those in urban areas. Only OALWH who were on long-term HIV treatment were interviewed in this study; thus, their circumstances may also differ from those who are not in care or the newly diagnosed. Relatedly, there may be a survivor effect in the study population where OALWH who are most sick are unable to seek care, hence not part of the studied population. As is the norm for qualitative studies in general, data collection, analysis, and interpretation are subject to individual influences; nonetheless, we countered this effect by maintaining reflexivity and constant discussion with the research team to provide rigour and credibility to the study.

## Conclusions

Our findings provide initial insight into the biopsychosocial challenges confronted by OALWH in a low-literacy Kenyan setting. The participants’ views indicate that mental complaints (especially common mental disorders and memory difficulties), physical problems (particularly comorbidities and somatic symptoms) and psychosocial challenges (especially poverty, stigma, and discrimination) are of concern among OALWH. Many of the perceived risk factors for these challenges often overlap across the biopsychosocial domains. Our study also highlights several opportunities for interventions and future research to tackle these issues in the study setting. Most significantly, any intervention response should be cohesive and collaborative to achieve maximum benefits and target modifiable factors and consider developing and nurturing the coping strategies of OALWH to aid self-care. Before programmes can have any impact on OALWH, improved access to basic needs, including food, financial support, and caregiving, and stigma and discrimination need to be addressed. Future research should quantify the burden of these challenges, examine the resources available to these adults, pilot, and test feasible interventions in this setting; and in doing so, aim to improve the lives of older adults living with HIV.

## Data Availability

No additional data available. The data that support the findings of this study have been included in this article.

## Acknowledgements

We would like to thank all the participants who took part in this study. We also acknowledge Irene Kasichana, Maureen Nyadzua, Haprity Mwangata, Linda Moranga and Victor Mwalewa for their role in data transcription. This work is published with the permission of the director of Kenya Medical Research Institute.

